# Multilingual Evaluation of a Large Language Model-Based Primary Care Chatbot

**DOI:** 10.64898/2026.05.03.26352241

**Authors:** Pei-Lun Chen, Amogh Ananda Rao, Sydney Pugh, Kevin B Johnson

## Abstract

Pre-visit planning has the potential to reduce EHR documentation burden while improving workflow efficiency, care quality, and patient–provider engagement. Large language model (LLM) chatbots show promise for supporting this task, but while their English-centric development suggests a potential for disparity, the extent to which these concerns translate into performance degradation in multilingual clinical settings remains unclear. In this mixed-methods study, we systematically evaluate the multilingual capabilities of PCP-Bot, an English-developed LLM-based (GPT-4o) clinical chatbot that collects patient concerns and generates structured, physician-ready summaries (∼200 words) under structured output constraints. We enrolled 31 bilingual individuals (11 Mandarin, 10 Spanish, 10 Hindi) to role-play as patients to evaluate the PCP-Bot, interacting with it across five synthetic clinical cases in both English and a second language. Participants completed a structured survey comprising baseline language proficiency screening, standardized interactions with PCP-Bot in each language, and post-interaction evaluations. Case order was randomized, with each scenario completed first in English and subsequently in the participant’s second language. All summaries were generated in English, regardless of the interaction language. Our results show that Hindi achieved usability and conversation quality parity with English across all measured dimensions. Mandarin achieved usability parity but showed a significant conversation quality gap relative to English. Spanish demonstrated significant deficits in both conversation quality and summary quality. Trust and workload remained consistent across languages. Qualitatively, participants found PCP-Bot natural, smooth, and accurate overall, but noted repetition, transcription errors, missed follow-ups, and more frequent usability issues in non-English interactions. Overall, our findings demonstrate that LLM translation capabilities can enable effective deployment beyond English following appropriate performance validation.

## Introduction

Electronic health records (EHRs) have transformed medical documentation by enhancing data accessibility and continuity of care. However, this digital shift has also introduced significant administrative demands on physicians, with observational and survey-based studies indicating that 35% to 37% of their time during patient encounters is devoted to EHR-related tasks^1,2^. This increasing administrative load not only affects clinical efficiency and physician well-being but also reduces the time available for direct, meaningful interactions with their patients. To help mitigate these challenges, strategies such as pre-visit planning have emerged^1^. Pre-visit planning ensures providers review essential patient information and medical history before the encounter, optimizing healthcare delivery by enabling more efficient visits, improved health outcomes, and enhanced patient satisfaction through timely, informed decision-making^3^.

Medical chatbots have emerged as a promising solution to streamline clinical documentation, reduce administrative burden, and enhance physician-patient interactions by automating routine tasks and supporting real-time data entry^2,3^. Many of these chatbots use large language models (LLMs), which are AI systems trained on vast amounts of text by leveraging deep learning methods to recognize complex word relationships and perform natural language processing tasks^4^.

Unlike traditional rule-based chatbots, LLMs have the capability of communicating in hundreds of languages. This feature of LLM-based chatbots has the potential to transform the communication between caregivers and the more than 29 million limited English proficiency (LEP) individuals in the United States^4^. These individuals face significant healthcare disparities, lower rates of preventive care and poorer health outcomes despite current government efforts^5^. A 2012 study found that patients with LEP who did not receive professional interpretation at admission had hospital stays 0.75–1.47 days longer and higher 30-day readmission rates^6^. Efforts to address this rely on a fragmented mix of professional interpreters, remote video or phone services, and untrained informal translators such as family members^7^. This fragmentation is driven in part by limited financial support, as most third-party payers, including Medicaid, do not reimburse for language services^7^. These approaches remain constrained by interpreter shortages, equipment failures, and inconsistent protocols^5^. Recent advances in LLM-based communication tools may address these gaps.

LLMs show promise in simplifying medical information and improving health literacy, offering a scalable, 24/7 approach to clinical communication^9^. However, this field remains early in its validation^8,9^ and performance is not uniform across populations. For example, GPT-3.5 was shown to be 5.82 times more likely to produce incorrect responses in non-English languages, with performance declines of 9.1% in Spanish, 28.3% in Chinese, and 50.5% in Hindi^9^. These disparities were partly attributed to subordinate bilingualism, in which models translate inputs into English before generating outputs, introducing errors in language-specific contexts^10^. Newer models such as GPT-4 have improved multilingual and multimodal performance. Studies demonstrate high accuracy (98–100%) in extracting explicit clinical information across languages, but reduced performance on tasks requiring contextual inference^10^. Similarly, GPT-4o has shown promise in real-time interpretation tasks in clinical settings^11^.

Despite these advances, most evaluations focus on structured or controlled tasks. Little is known about how LLMs perform in real-world, multilingual clinical workflows, especially in supporting care for patients with limited English proficiency, where communication barriers contribute to disparities^12^. To address this gap, we evaluated a pre-visit chatbot (PCP-Bot), an LLM-based system built on GPT-4o that collects patient concerns and generates structured, physician-ready summaries under strict guardrails prohibiting medical advice or diagnostic claims^13^. In this study, we conducted a multilingual study assessing its performance across English, Mandarin Chinese, Spanish, and Hindi, the four most widely spoken global languages^14^.

## Methods

### 1. Study Design

We conducted a mixed-methods study in which bilingual participants simulated patient interactions with PCP-Bot^12^ using five randomly presented, synthetic clinical cases, first in English then a second language (Mandarin Chinese, Spanish, Hindi). The experimental workflow is visualized in **Figure 1**.

**Figure 1.**
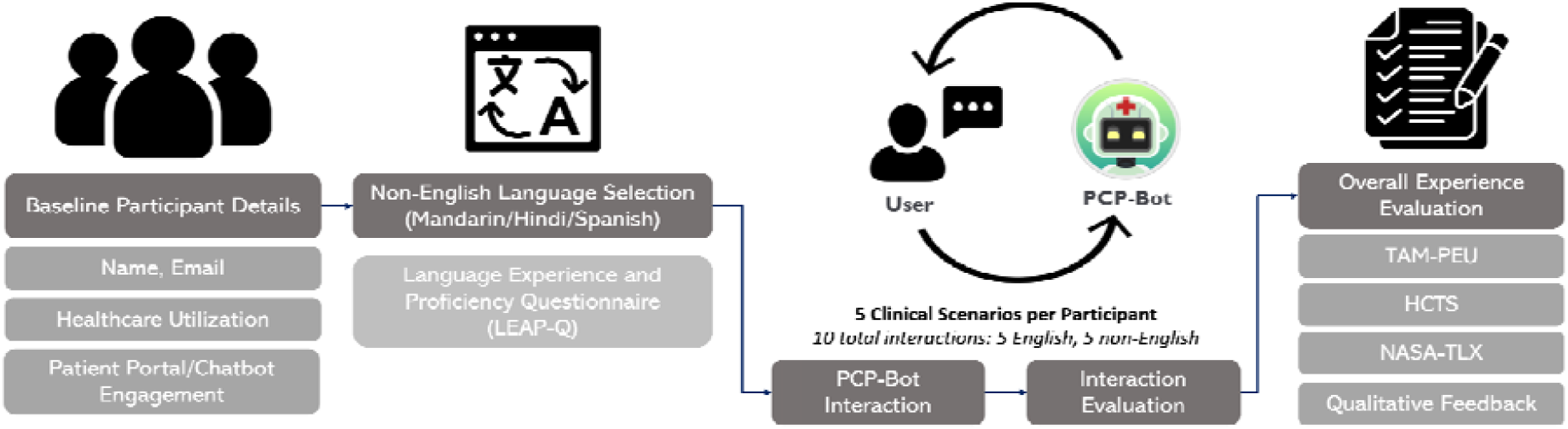
Overview of the PCP-Bot Multilingual Study Workflow and Evaluation Framework

### 2. Study Instruments

#### 2.1 PCP-Bot: Conversational Pre-Visit Assistant

PCP-Bot is a voice-based conversational assistant designed to conduct pre-visit intake interviews. Participants reviewed each synthetic clinical case and then engaged with PCP-Bot in the role of the described patient. At the conclusion of each interaction, PCP-Bot generated a structured summary of the gathered information. For non-English interactions, summaries were produced in English to enable standardized review across languages. System architecture is illustrated in Figure 2.

**Figure 2.**
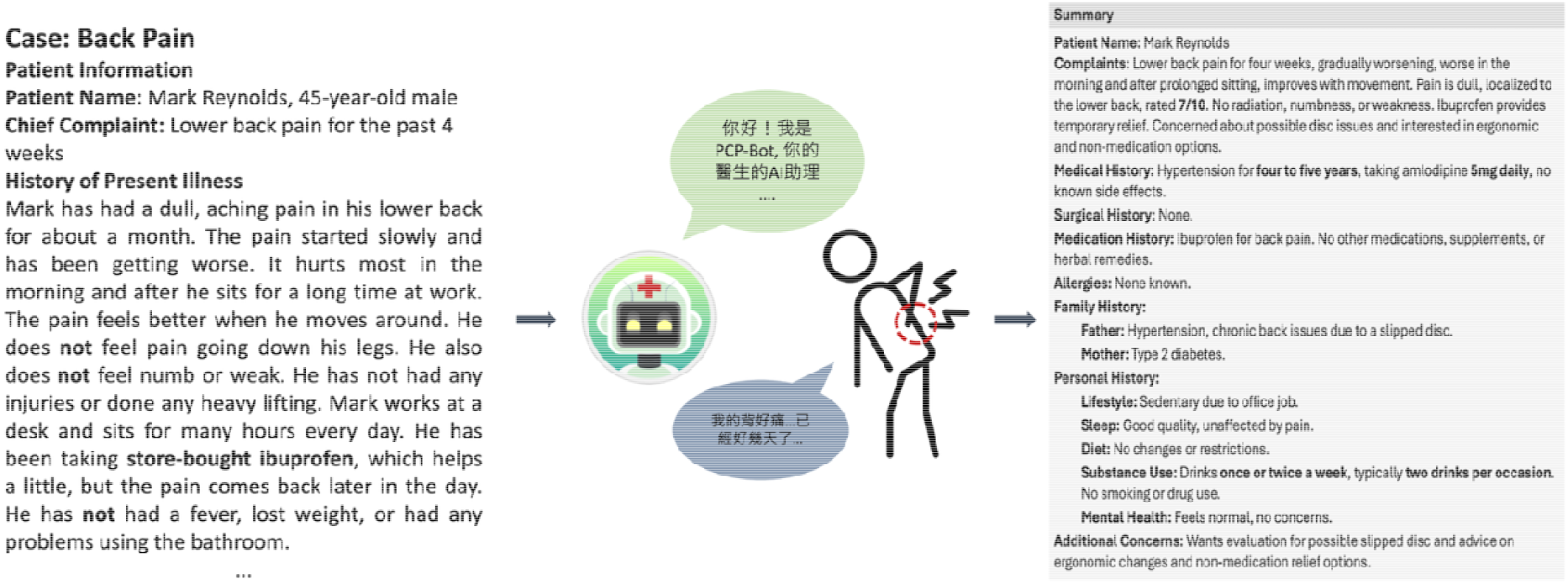
Example of an Interaction between a Patient and PCP-Bot

#### 2.2 Synthetic Clinical Cases

Six synthetic clinical cases were developed to represent typical primary care chief complaints, spanning gastrointestinal, musculoskeletal, urinary, neurological, metabolic, and respiratory presentations. Each case included relevant medical history, lifestyle factors, and patient concerns. Five cases (C1–C5) were used in the evaluation; one additional case (C0) served as a familiarization exercise prior to the study interactions. Case details are summarized in Table 1; the full C0 scenario is provided in Appendix A.

**Table 1.**
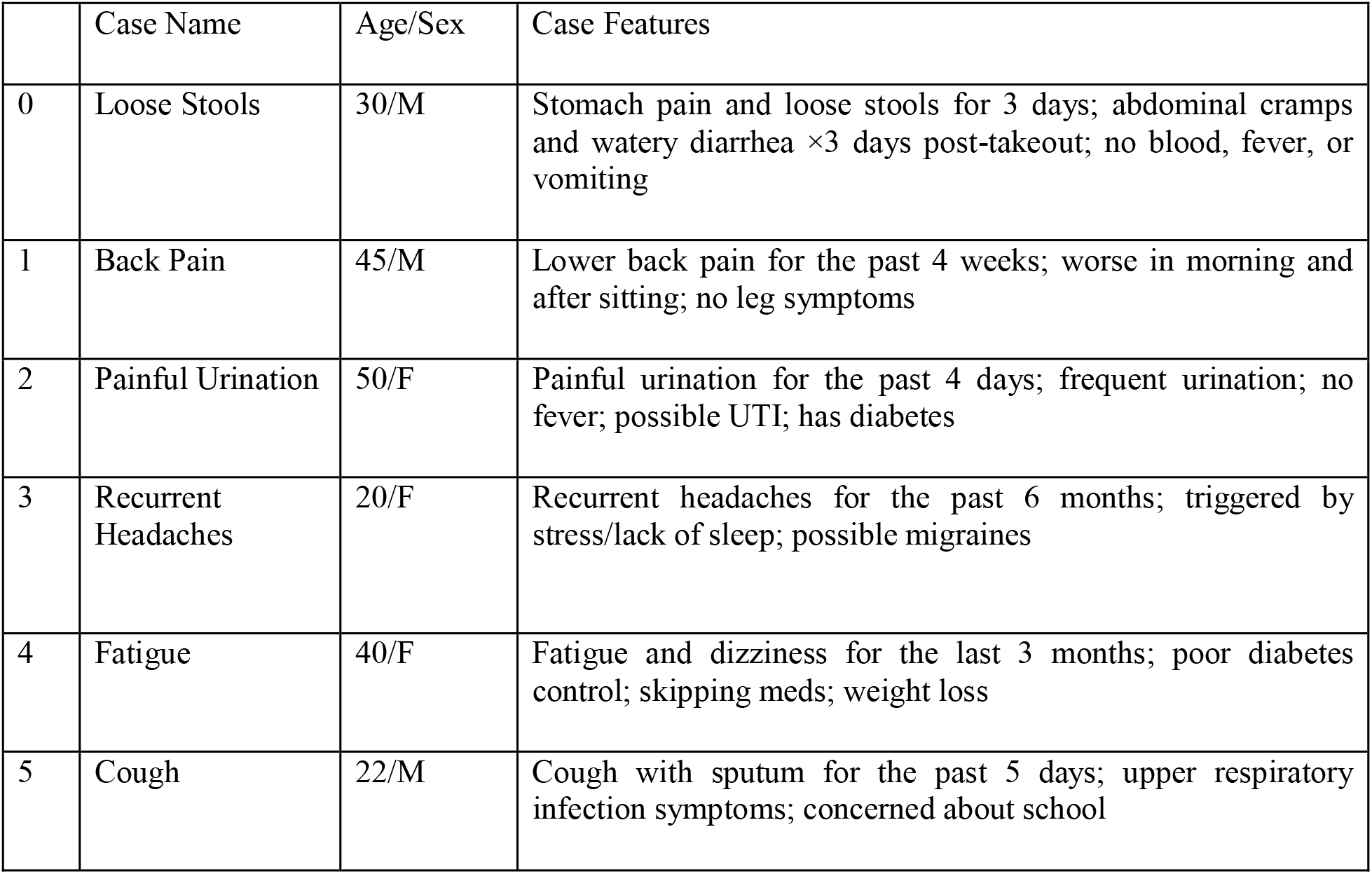
Summary of Standardized Patient Cases.

|  | Case Name | Age/Sex | Case Features |
| --- | --- | --- | --- |
| 0 | Loose Stools | 30/M | Stomach pain and loose stools for 3 days; abdominal cramps and watery diarrhea ×3 days post-takeout; no blood, fever, or vomiting |
| 1 | Back Pain | 45/M | Lower back pain for the past 4 weeks; worse in morning and after sitting; no leg symptoms |
| 2 | Painful Urination | 50/F | Painful urination for the past 4 days; frequent urination; no fever; possible UTI; has diabetes |
| 3 | Recurrent Headaches | 20/F | Recurrent headaches for the past 6 months; triggered by stress/lack of sleep; possible migraines |
| 4 | Fatigue | 40/F | Fatigue and dizziness for the last 3 months; poor diabetes control; skipping meds; weight loss |
| 5 | Cough | 22/M | Cough with sputum for the past 5 days; upper respiratory infection symptoms; concerned about school |

#### 2.3 PCP-Bot Evaluation Survey

The survey comprised three phases: Baseline Assessment, Scenario-Based Interaction, and Overall Subjective Experience. During the Baseline Assessment, participants provided demographic information and completed questionnaires on self-reported language proficiency (speaking, comprehension, reading), prior healthcare utilization, digital health tool engagement, and chatbot experience. Participants then selected their target non-English evaluation language and identified their primary language by indicating which language dominated their lifetime environment.

We used a modified version of the LEAP-Q^15,16^ (Appendix B) survey for language proficiency. This survey uses self-ratings (0–10) across speaking, comprehension, and reading; primary language proficiency was assumed perfect (score = 10). Precise characterization of linguistic ability was not an objective of this study; these ratings served to establish an evaluator baseline rather than to assess language competence per se.

During the Scenario-Based Interaction phase, participants completed all five cases twice (once in English and once in their non-English language) with case order randomized to minimize carryover effects. After each interaction, participants completed the Bot Usability Scale (BUS-11)^17,18^, a validated 11-item instrument measuring chatbot interaction quality across five factors: Accessibility, Functionality, Conversation, Privacy, and Responsiveness (5-point Likert scale; 1 = strongly disagree, 5 = strongly agree) (Appendix C). Participants also rated overall communication quality on a 0–5 scale (0 = extremely poor, 5 = perfect). Following each interaction, participants evaluated the PCP-Bot-generated summary using an adapted rubric based on Khanna and colleagues^19^, with items modified to fit the study context. Summaries were rated on four criteria: Fluency (readability and grammatical accuracy), Adequacy (information preservation), Meaning (semantic equivalence and misinformation detection), and Severity (potential impact on patient health outcomes), each on a 5-point Likert scale (Appendix D)^19^.

The Overall Subjective Experience phase used three validated instruments: TAM-PEU^20,21^, which measures perceived ease of use as a predictor of technology adoption; HCTS^22,23^, which assesses trust in technology as a socio-technical construct; and NASA-TLX^24^, which measures subjective workload across six dimensions (mental, physical, and temporal demand; effort; performance; frustration). Items are provided in Appendices E–G.

Qualitative feedback was collected via open-ended free-text responses embedded after each interaction and at the survey’s conclusion. Participants were prompted to describe difficulties communicating with PCP-Bot, perceived differences between English and non-English interactions, overall challenges encountered, and expectations for AI-powered clinical chatbots. Responses were optional; participants without issues were instructed to enter “N/A.”

### 3. Testing Protocol

#### 3.1 Participant Recruitment

The study population consisted of bilingual individuals who volunteered to test PCP-Bot and annotate its generated pre-visit summaries. Eligible participants self-reported fluency in English and one of the evaluated non-English languages (Mandarin Chinese, Hindi, or Spanish). We aimed to recruit thirty participants, ten for each target non-English language.

#### 3.2 Participant Interaction Procedure

Participants first attended an information session, during which a brief demonstration of PCP-Bot was provided along with visual aids to facilitate use. Following the session, participants completed the Qualtrics survey asynchronously at their own convenience. PCP-Bot was accessed directly from within the survey via a QR code, and all interactions were conducted on participants’ personal mobile devices. After completing each interaction, participants returned to the survey to complete the corresponding evaluation measures. For each interaction, participants were required to paste both the link to their PCP-Bot conversation and the generated summary into the survey to enable verification and analysis. For non-English interactions, participants were instructed to ask PCP-Bot for an additional English summary following interaction. Participants were able to pause and resume the survey at any time, allowing flexible completion across multiple sessions. Each participant received a $50 USD gift card after submitting the completed survey.

### 4. Ethical Considerations

This study was reviewed and deemed exempt by the Institutional Review Board of the University of Pennsylvania Human Research Protections Program (HRPP). No real patient data was used, and all interaction data were de-identified and securely stored.

### 5. Funding Declaration

This work was supported by the National Library of Medicine and the NIH Office of the Director (Project No. 5DP1LM014558-03; former No. 1DP1OD035237-01).

### 6. Data Analysis

Data cleaning and preparation were performed in Python (version 3.9.6). Incomplete entries and non-English AI-generated summaries were excluded; corresponding Summary Evaluation scores were removed to ensure consistency. Likert-scale responses were encoded as ordinal integers (1–5). Composite scores for BUS-11, Summary Quality, NASA-TLX, HCTS, and TAM-PEU were computed using established scoring conventions. NASA-TLX items were reverse-coded and rescaled to a 0–100 range; Summary Quality Severity N/A responses were encoded as 0. BUS-11, Summary Quality, and Conversation Quality scores were computed per interaction and averaged across five cases (C1–C5), yielding one composite score per participant per measure.

Statistical analyses were conducted in RStudio (version 4.5.0). Normality was assessed using the Shapiro–Wilk test alongside skewness and kurtosis evaluation; given non-normal distributions, Mann– Whitney U tests were used to compare outcomes between English and non-English interactions, with Kruskal–Wallis tests and Wilcoxon post hoc comparisons applied for differences across language groups. Descriptive statistics (mean, SD, median) were calculated for all quantitative outcomes. Qualitative data were reviewed independently by team members; recurring patterns were synthesized into descriptive themes through discussion and consensus.

## Results

### Language Proficiency

We enrolled 31 bilingual individual participants (ages 20 – 30): 11 evaluating Mandarin Chinese, 10 Spanish, and 10 Hindi. As shown in Figure 2, self-reported proficiency was uniformly high across all evaluated languages and skills (speaking, comprehension and reading.) with mean scores ranging from 8.81 to 9.82 across groups and domains. No significant differences were observed between language groups on any proficiency dimension (Kruskal–Wallis, all p > 0.37).

Primary language alignment varied by group: 81.8% of Mandarin evaluators identified Mandarin as their primary language, compared to 60% for Spanish and 50% for Hindi (Table 2). Despite this variation, proficiency ratings remained high across all groups, suggesting participants were confident and capable in their evaluation language regardless of primary language dominance.

**Table 2.** Self-Reported Language Proficiency by Language and Skill (Mean ± SD)

| Skill | English (n=31) | Mandarin Chinese (n=11) | Spanish (n=10) | Hindi (n=10) |
| --- | --- | --- | --- | --- |
| Speaking | 8.81 ± 1.80 | 9.55 ± 1.04 | 9.20 ± 1.32 | 8.90 ± 2.28 |
| Comprehension | 9.13 ± 1.41 | 9.82 ± 0.60 | 9.40 ± 1.07 | 9.40 ± 1.07 |
| Reading | 9.16 ± 1.32 | 9.64 ± 0.92 | 9.5 ± 0.71 | 9.40 ± 1.26 |

*No significant differences between language groups on any proficiency dimension (Kruskal–Wallis, all $p > 0.27$ )*

### Healthcare & Health Technology Use

Participants were regular healthcare users: 48.4% reported 1–2 visits in the past year and 45.2% reported 3–4. Engagement with digital health tools was lower. Approximately half reported never using health apps or patient portals in the past month. Most participants had prior chatbot experience (90.3%, n = 28), providing a relevant baseline for evaluating PCP-Bot usability. Language-specific patterns are summarized in Table 3.

**Table 3.** Participant Healthcare Utilization and Digital Tool Engagement by Language Group.

| Language | Response Category | Healthcare Visits (past year) | Health App Usage (past month) | Patient Portal Usage (past month) | Previous Chatbot Usage |
| --- | --- | --- | --- | --- | --- |
| Overall (n=31) | Never | 3.2% | 48.4% | 41.9% | Yes: 90.3%<br>No: 9.7% |
|  | Rarely (1-2×) | 48.4% | 41.9% | 45.2% |  |
|  | Sometimes (3-4×) | 45.2% |  |  |  |
|  | Often (5-7x) | 3.2% | — | — |  |
|  | Very Often (> 8x) | — | — | 3.2% |  |
| By language group |  |  |  |  |  |
| Mandarin Chinese (n=11) | Never | — | 63.6% | 63.6% | Yes: 81.8%<br>No: 18.2% |
|  | Rarely (1-2×) | 63.6% | 27.3% | 27.3% |  |
|  | Sometimes (3-4×) | 27.3% | 9.1% | 9.1% |  |
|  | Often (5-7x) | 9.1% | — | — |  |
|  | Very Often (> 8x) | — | — | — |  |
| Spanish (n=10) | Never | — | 50.0% | 20.0% | Yes: 90.0%<br>No: 10.0% |
|  | Rarely (1-2×) | 30.0% | 40.0% | 50.0% |  |
|  | Sometimes (3-4×) | 70.0% | 10.0% | 20.0% |  |
|  | Often (5-7x) | — | — | — |  |
|  | Very Often (> 8x) | — | — | 10% |  |
| Hindi (n=10) | Never | 10.0% | 30.0% | 40.0% | Yes: 100%<br>No: 0% |
|  | Rarely (1-2×) | 50.0% | 60.0% | 60.0% |  |
|  | Sometimes (3-4×) | 40.0% | 10.0% | — |  |
|  | Often (5-7x) | — | — | — |  |
|  | Very Often (> 8x) | — | — | — |  |

— indicates 0% (no participants in this response category). Chatbot experience is shown once per group (yes/no dichotomy)

### Chatbot Usability Scale (BUS-11)

Perceived usability was high across all languages, with mean BUS-11 scores of 4.54 for English, 4.33 for Mandarin Chinese, 4.45 for Spanish, and 4.42 for Hindi (non-English mean: 4.40; Table 4; Figure 3A). No significant difference was found between English and non-English interactions overall (Mann-Whitney U, p = 0.128), and pairwise comparisons confirmed no significant differences for any individual language pair (English vs. Mandarin Chinese: p = 0.18; English vs. Spanish: p = 0.29; English vs. Hindi: p = 0.43).

**Table 4.** Objective and Subjective Performance of PCP-Bot Interactions Across Languages.

| Measure / Test | English | Non-English |  |  |
| --- | --- | --- | --- | --- |
|  |  | Mandarin Chinese | Spanish | Hindi |
| BUS-11 (usability) | 4.54 | 4.33<br>($p = 0.18$) | 4.45<br>($p = 0.29$) | 4.42<br>($p = 0.43$) |
| Conversation Quality | 4.42 | 3.89<br>($p < 0.05$) | 3.56<br>($p < 0.05$) | 3.98<br>($p = 0.16$) |
| Summary Quality | 3.88 | 3.72<br>($p = 0.05$) | 3.60<br>($p < 0.05$) | 3.67<br>($p = 0.44$) |
| TAM-PEU (ease of use) | - | 5.92<br>($p = 0.13$) | 6.25<br>($p = 0.13$) | 6.40<br>($p = 0.13$) |
| HCTS (trust) | - | 4.54<br>($p = 0.26$) | 4.87<br>($p = 0.26$) | 5.04<br>($p = 0.26$) |
| NASA-TLX (workload) | - | 41.5<br>($p = 0.30$) | 31.7<br>($p = 0.30$) | 44<br>($p = 0.30$) |

*P* values reflect pairwise comparisons (BUS-11, conversation quality, summary quality), and Kruskal–Wallis tests (TAM-PEU, HCTS, NASA-TLX)

**Figure 3.**
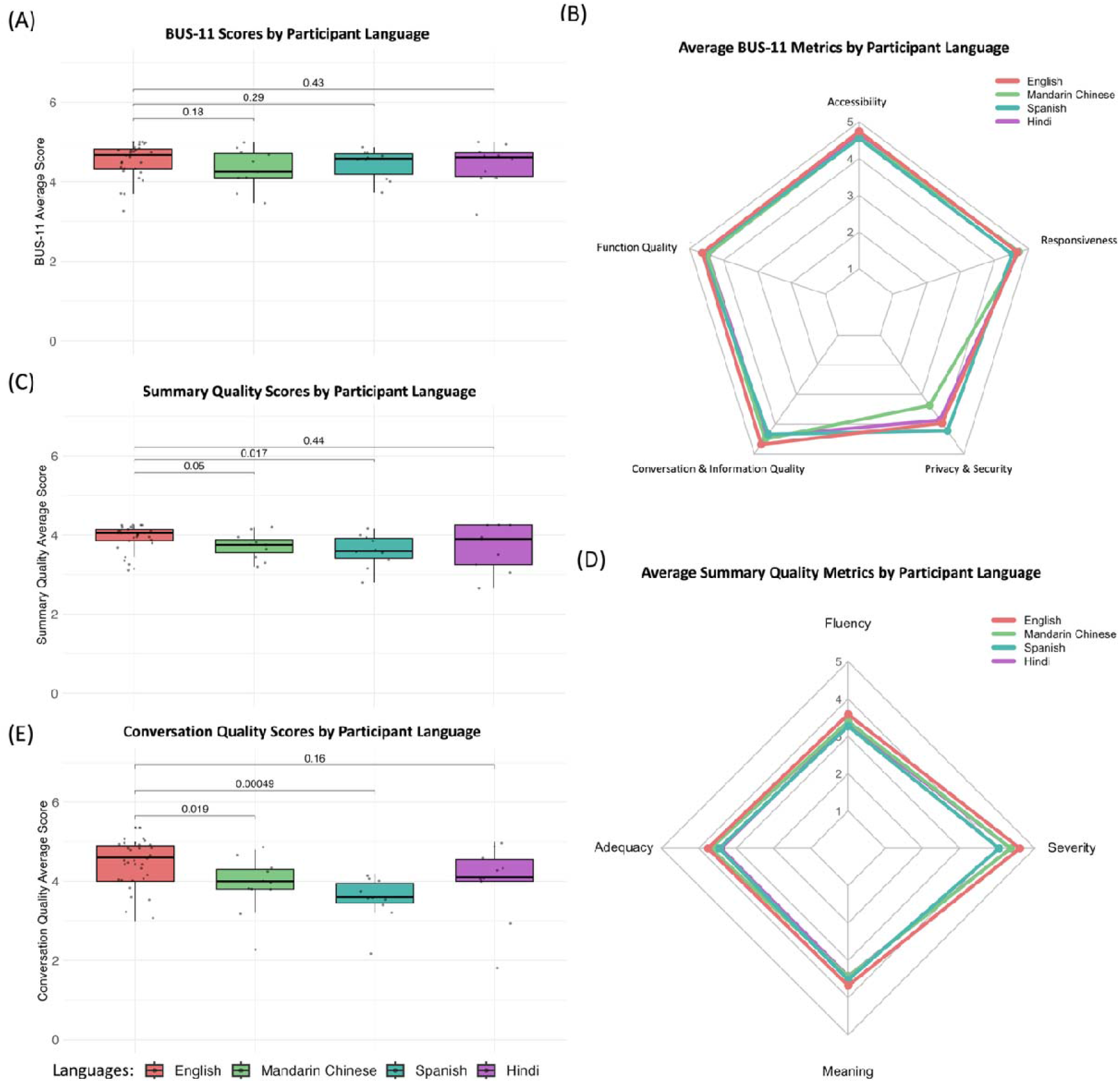
Quantitative Assessment of PCP-Bot Performance Across Languages: (A) User-Reported Usability (BUS-11) (B) BUS-11 Metric Distribution by Language (C) Summary Quality Score (D) Summary Quality Metric Distribution by Language (E) Conversation Quality Score

Decomposition into BUS-11 component subscales (accessibility, responsiveness, privacy and security, conversation and information quality, and function quality) showed that English scored highest across all categories, with non-English languages exhibiting slightly lower but consistent profiles (Figure 3B). Ratings were consistent across languages for accessibility, responsiveness, and function quality, with slightly more variation in privacy & security and conversation & information quality.

### Conversation Quality

Conversation quality scores showed clear cross-lingual differences, with means of 4.42 for English, 3.89 for Mandarin Chinese, 3.56 for Spanish, and 3.98 for Hindi (non-English mean: 3.81; Table 4; Figure 3E). A Mann-Whitney U test confirmed a statistically significant difference between English and non-English interactions (p < 0.001). Pairwise comparisons identified significant gaps for Spanish (p < 0.001) and Mandarin Chinese (p = 0.019) relative to English, with Spanish showing the larger deficit; Hindi did not differ significantly from English (p = 0.16).

### Summary Quality

Of 310 recorded summaries, incomplete entries and non-English-language outputs were excluded. One participant whose non-English summaries were not generated in English was excluded entirely from Summary Quality analyses, as their scores could not be averaged across cases. The retained sample included 30 English, 11 Mandarin Chinese, 10 Spanish, and 9 Hindi summaries.

Summary quality scores were modest but consistently above the scale midpoint across all languages (Figure 3C), with means of 3.88 for English, 3.72 for Mandarin Chinese, 3.60 for Spanish, and 3.67 for Hindi (non-English mean: 3.66; Table 4). A Mann-Whitney U test confirmed a statistically significant difference between English and non-English interactions (p = 0.0137). Pairwise comparisons identified significant gaps for Spanish (p = 0.017) and Mandarin Chinese (p = 0.05) relative to English; Hindi did not differ significantly (p = 0.44).

Analysis of individual dimensions (Fluency, Adequacy, Meaning preservation, and Severity) showed English interactions achieving the highest scores on the first three (Figure 3D). Non-English interactions produced slightly lower fluency, adequacy, and meaning preservation scores alongside higher Severity ratings, indicating that errors in non-English summaries carried greater potential impact on patient health outcomes.

### Overall Subjective Experience Metrics

TAM-PEU scores were high across all non-English language groups, with no statistically significant differences between them (Table 4). Overall trust measured by the HCTS was similarly moderately high across languages, with no significant differences (Table 4). Examination of HCTS subdimensions revealed consistently high perceived competency, general trust, benevolence, and reciprocity alongside relatively low risk perception, indicating that users generally did not feel unsafe, vulnerable, or cautious when interacting with the system. Hindi users consistently reported the highest trust-related scores, while Mandarin users showed slightly lower values across several dimensions. Workload assessed via NASA-TLX further supported these findings, with low perceived demand across all language groups and no significant differences (Table 4). Radar plots showed uniformly low workload and frustration across all NASA-TLX dimensions (Figure 4), with only two isolated outliers (one Mandarin, one Hindi) reporting very high effort (80/100).

**Figure 4.**
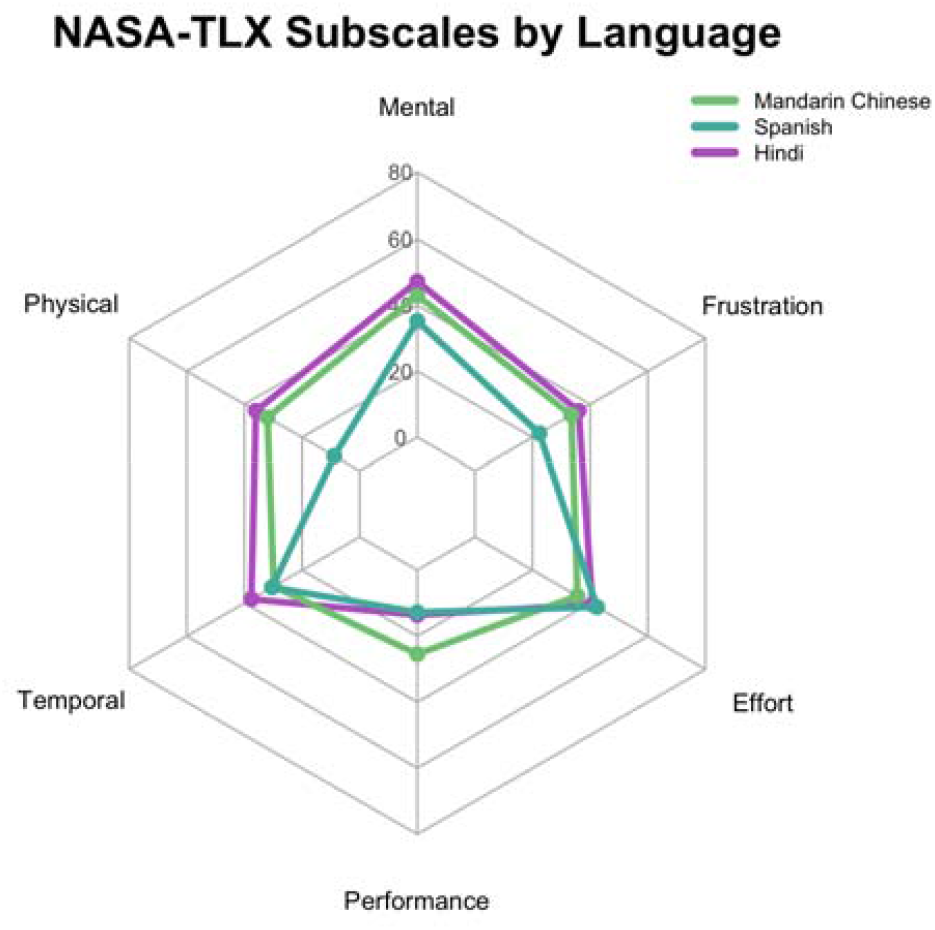
NASA-TLX Metric Bucket Distribution by Language

### Qualitative Results

Participants generally described PCP-Bot as natural, smooth, and accurate, with several noting that non-English interactions performed better than expected. One demonstrated promising capability, with the non-English version capturing “almost all the details from the beginning” and touching “on every section,” and one participant observing that “the conversation in Mandarin required less prompting from me to receive all necessary information”. Where translation fell short, participants noted that the system often remained functionally intelligible; as one put it, the bot sometimes had difficulty “distinguishing between Hindi and Urdu, but understands the overall gist of the answer, so does not affect the responses as much,” and another remarked that “the English summary translation from Hindi could be better, but overall message is conveyed well”.

Recurring limitations included repetition across languages (the bot was described as “very repetitive,” particularly in Hindi), transcription errors (misidentifying names, misinterpreting clinical details such as “yellow sputum”), and insufficient follow-up questioning. One participant noted the bot “was not asking any follow-up questions, so I had to provide all the information,” and another observed it follows up in English “but not in Hindi.” Additional issues specific to non-English interactions included mid-conversation reversion to English, poor spoken Spanish quality, and a faster speech rate that participants found harder to understand.

## Discussion

In this study, we evaluated the multilingual capabilities of the English-developed ChatGPT-based pre-visit chatbot using a mixed-methods design where bilingual participants interacted with the system within a standardized survey environment and subsequently provided quantitative and qualitative evaluations of usability, trust, workload, and overall experience. Overall, our results show that PCP-Bot demonstrates strong cross-lingual generalizability, with consistently high usability, functionality, and perceived usefulness across English, Spanish, Hindi, and Mandarin Chinese.

One strength of this study is the high linguistic proficiency of the participant cohort. Participants reported near-native proficiency across all evaluated languages, with no statistically significant differences between language groups. This effectively controls user-side language barriers, supporting strong internal validity. As a result, observed differences in performance can be attributed primarily to the chatbot itself rather than variability in user comprehension or communication ability. Participant technology profiles show that most have had extensive prior experience with chat-based interfaces. However, engagement with healthcare-specific digital tools (i.e. patient portals, health apps) was relatively low and consistent across language groups. This suggests that favorable evaluations of PCP-Bot were not driven by prior familiarity with healthcare technology but rather reflect the system’s inherent usability and accessibility.

Across core performance metrics, PCP-Bot demonstrated high usability and technical robustness in all languages. BUS-11 scores were uniformly high with no statistically significant pairwise differences across any language group, indicating that the system is accessible, responsive, and easy to use regardless of language. Notably, usability subdomains related to accessibility and responsiveness were consistent across languages, while greater variability emerged in domains tied to more human-like qualities, such as conversational flow and perceived privacy or security. This pattern suggests that system-level functionality translates well across languages, while more nuanced aspects of interaction remain sensitive to linguistic and cultural differences. Conversational quality showed modest but meaningful cross-lingual variation. While Hindi interactions achieved near–English-level performance with no statistically significant difference (p = 0.16), both Mandarin Chinese (p = 0.019) and Spanish (p = 0.00049) demonstrated significant performance gaps relative to English, with Spanish showing the larger deficit. This suggests that PCP-Bot’s cross-lingual capabilities are uneven across languages and that Spanish and Mandarin Chinese functionality may benefit from targeted improvement.

Despite these differences, overall scores remained high, and distributions overlapped substantially across languages, indicating that the chatbot can sustain generally coherent and effective interactions, with residual gaps likely reflecting differences in linguistic nuance, contextual phrasing, or cultural alignment. While summary quality showed modest cross-lingual variation with a significant reduction for Spanish (p = 0.017) and a marginal difference for Mandarin Chinese (p = 0.05), Hindi was fully comparable to English (p = 0.44), and all scores remained above the scale midpoint, suggesting generally effective summarization across languages. The chatbot’s ability to extract, synthesize, and structure clinically relevant information remained consistent regardless of input language. This suggests that the underlying information processing and summarization pipeline is robust to linguistic variation, even when surface-level conversational differences exist. From a clinical perspective, this is a critical finding, as it indicates that multilingual inputs can be reliably transformed into standardized, physician-ready outputs.

The Spanish performance deficit likely reflects compounding technical and data-related limitations. One potential reason for this deficit is the fundamentally heterogeneous nature of Spanish as a dialect continuum rather than a monolithic language^25^. Unlike languages with a single national standard, Spanish spans 21 countries with substantial regional lexical, morphological, and syntactic variation^25,26^. Studies evaluating LLM dialectal knowledge across Spanish-speaking nations has found that models reliably handle Castilian and Mexican varieties while performing substantially worse on Caribbean, Chilean, and Central American one^26^. Beyond vocabulary and syntax, cultural pragmatic differences introduce subtler but equally important failure modes. Spanish-speaking patients from certain cultural backgrounds may communicate health concerns indirectly, embed symptoms within personal narratives, or use idiomatic expressions to describe pain or distress that lack direct equivalents in standardized medical language^27^. A model trained predominantly on formal or clinical English text may fail to recognize these patterns, leading to incomplete extraction of clinically relevant information even when the conversational exchange appears superficially coherent.

When examining overall chatbot experience, perceived ease of use (TAM-PEU) was high across all groups, with no significant differences, indicating broad acceptability and willingness to adopt the tool. Trust (HCTS) was also consistently high, especially in domains such as reciprocity, where users reported a strong sense of cooperative interaction with the chatbot. However, slightly lower trust scores among Mandarin-speaking participants suggest that cultural or linguistic factors may influence perceptions of AI reliability and “personality.” Hofstede’s cultural dimensions theory suggests that in collectivist and high uncertainty avoidance cultures, which are prevalent across Asia, trust in AI is primarily driven by social influence and a collective reliance on shared norms to mitigate ambiguity^28,29^. China’s high power distance score (PDI = 80) and strong collectivism (IDV = 15) mean users likely evaluate AI through a lens of social hierarchy and group alignment rather than individual functional performance^28^. Workload assessments (NASA-TLX) indicated low perceived cognitive burden across all language groups. Results show that participants generally reported minimal effort, low frustration, and high task success, suggesting that the system is intuitive and does not introduce additional stress.

While English interactions yielded the highest overall scores, Mandarin Chinese and Hindi achieved usability and conversational performance comparable to English, suggesting that LLM-based clinical chatbots can effectively support multilingual workflows beyond English-centric settings. These results challenge the assumption that English-trained models are inherently limited in cross-lingual clinical applications and highlight the potential for broader deployment when appropriate safeguards and validation are in place.

Our work suggests LLM translation capabilities can support multilingual deployment and represent the initial step toward scalable cross-lingual generalization of clinical AI systems. However, multilingual deployment should be validated on a per-language basis to confirm performance consistency and safety across settings. Study limitations include our controlled, scenario-based design and relatively small participant cohort, which may limit generalizability to real-world clinical settings. Participants interacted with standardized, synthetic cases rather than presenting their own health concerns, which may not fully capture the complexity, variability, and emotional context of authentic patient–provider communication. In addition, the cohort was composed of highly proficient bilingual individuals, which, while strengthening internal validity, does not reflect the broader patient population. Summary quality was also evaluated by non-clinical assessors, which raises questions about the criteria used to determine whether all key clinical information was adequately captured.

Future work should extend evaluation to more diverse and representative patient populations, including individuals with heterogeneous language proficiency, health literacy, and access to digital technologies. Studies should move beyond controlled environments to assess real-world deployment within clinical workflows, where factors such as time constraints, competing demands, and integration with electronic health records may influence performance. Additionally, downstream outcomes such as clinical accuracy, documentation quality, clinician workload, clinic efficiency, and patient satisfaction should be assessed. Finally, longitudinal and implementation-focused studies are needed to ensure that multilingual deployment does not inadvertently exacerbate disparities but instead delivers equitable benefits across linguistic and cultural groups.

## Conclusion

This study evaluated a ChatGPT4o-based conversational pre-visit chatbot conversing in English, Mandarin Chinese, Spanish, and Hindi. Findings indicate high usability, trust, and low cognitive demand across all languages, with no significant differences on BUS-11 usability or perceived ease of use. Statistically significant gaps in conversation quality and summary accuracy emerged for Spanish and Mandarin Chinese, suggesting that GPT-based voice interaction and summarization are adequate, though not yet uniformly equivalent across languages. These findings are encouraging and suggest that focus on the gaps in fluency and meaning preservation for certain groups warrant attention before broader deployment.

## Data Availability

All data produced in the present study are available upon reasonable request to the authors

https://github.com/patrchen/PCP-Bot-Translator-Study-Cases

## Appendix

## APPENDIX A Standardized Synthetic Patient Cases for PCP-Bot Interaction (Case 0)

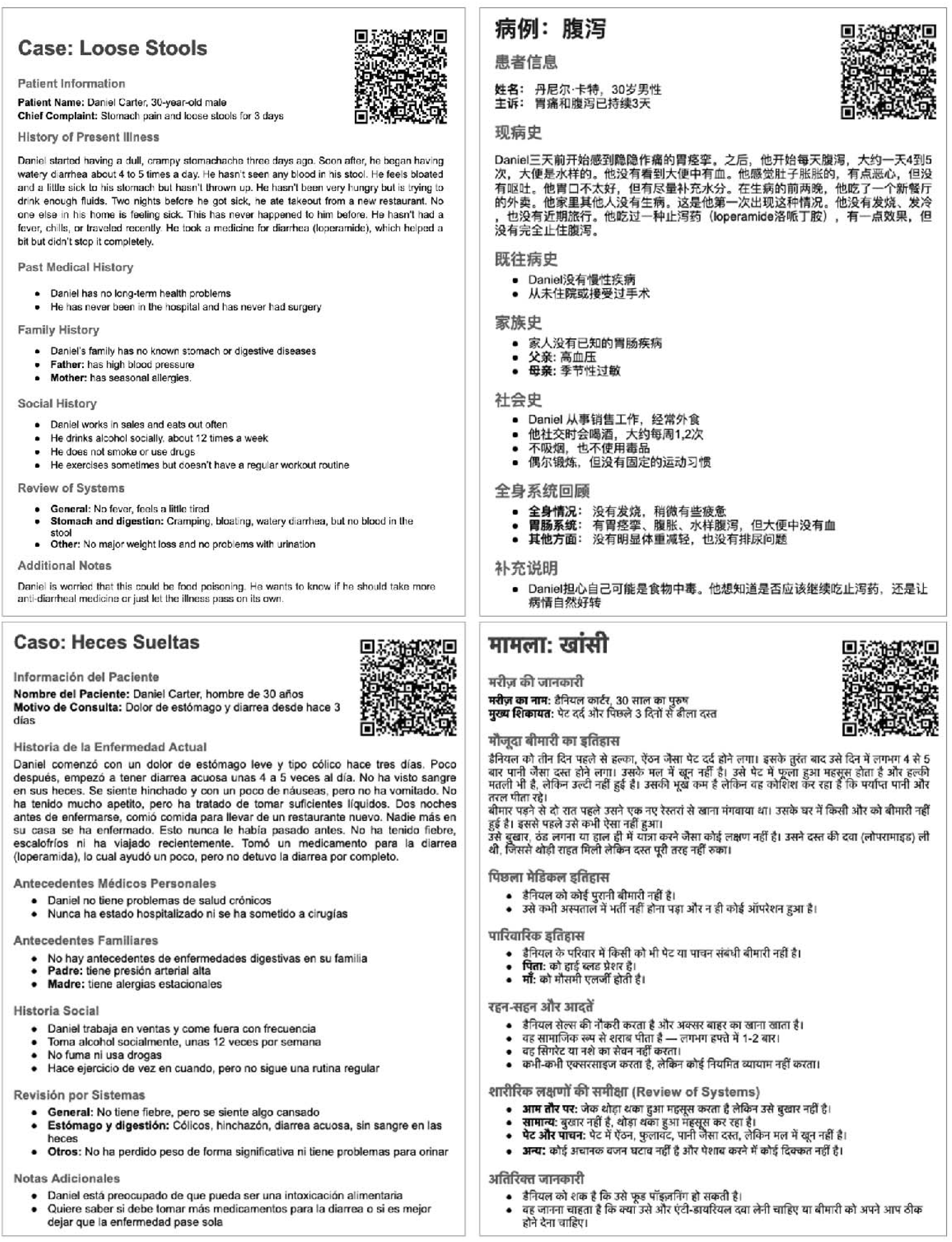

## APPENDIX B Adapted Baseline Language Proficiency Assessment Questionnaire

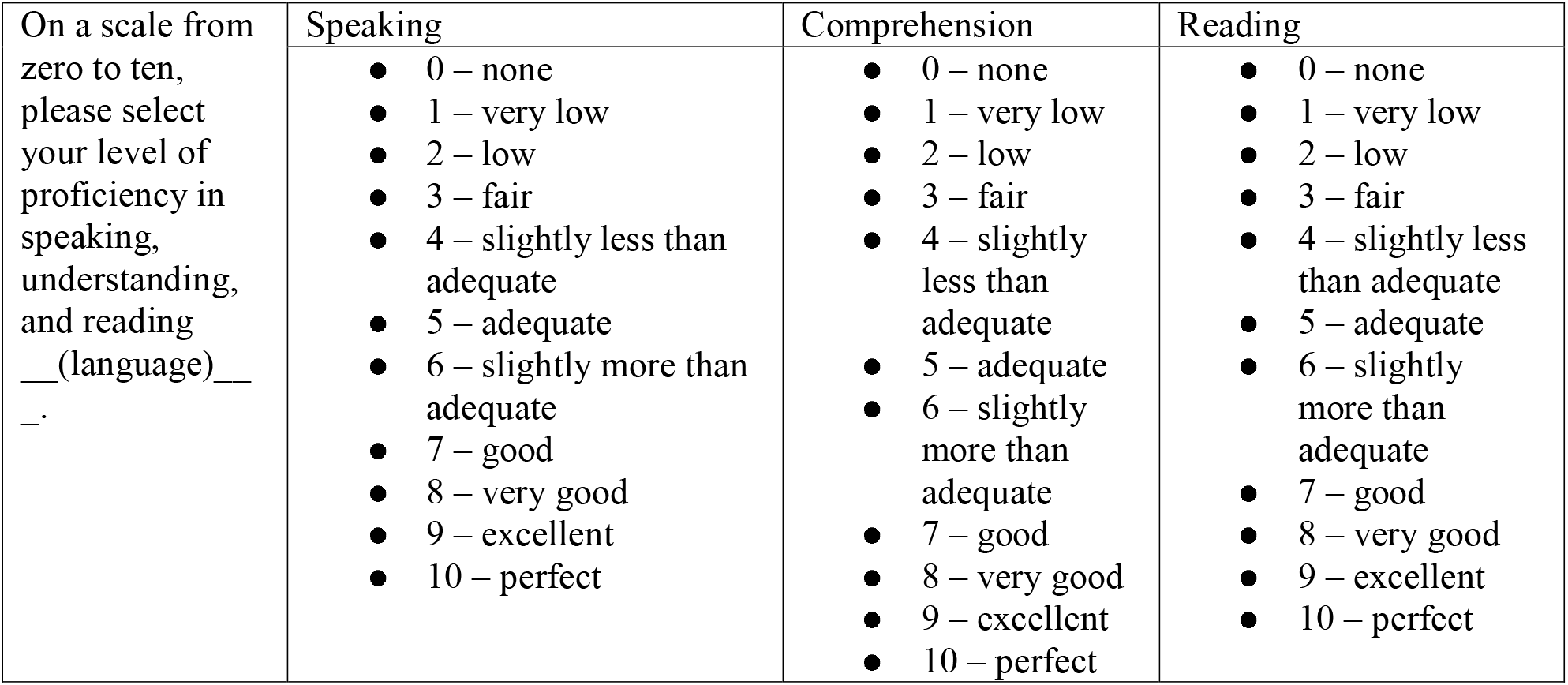

## APPENDIX C Adapted Chatbot Usability Scale (BUS-11) Questionnaire

Accessibility

- It was easy to find and use the features in PCP-Bot during this interaction
- It was easy to begin using PCP-Bot

Quality of Functions

- Communicating with PCP-Bot was clear and straightforward
- PCP-Bot was able to follow the flow of our conversation
- PCP-Bot’s responses were easy to understand

Quality of Conversation & Information

- PCP-Bot understood my responses and addressed them appropriately
- PCP-Bot gave an appropriate amount of information during the conversation
- PCP-Bot only provided information relevant to my medical history discussion
- PCP-Bot’s responses were accurate

Privacy & Security

- PCP-Bot made me aware of any possible privacy & security concerns

Responsiveness

- PCP-Bot responded quickly

## APPENDIX D Adapted Summary Quality Assessment Questionnaire

Fluency

How would you rate the fluency (grammar and readability) of the summary?

5 – Perfect Fluency; like reading a professionally written newspaper article

4 – Good fluency; minor grammar or style issues but easily understood

3 – Moderate fluency; noticeable grammar or style issues but still understandable

2 – Low fluency; difficult to read with significant grammar issues

1 – No fluency; no appreciable grammar, not understandable

Adequacy

How would you rate the adequacy of the summary in preserving the original information?

5 – 100% of information conveyed from the original conversation

4 – Most information preserved, minor omissions

3 – Some omissions or additions, but main ideas still present

2 – Significant omissions or additions; important details missing or altered

1 – 0% of information conveyed from the original

Meaning

How would you rate the meaning of the summary compared to the original conversation?

5 – Same meaning as the original

4 – Almost same meaning; very minor changes in nuance

3 – Some change in nuance, but main intent preserved

2 – Major change in connotation or intent

1 – Totally different meaning from the original

Severity

If there are errors, how severe could their impact be on the overall accuracy and usefulness of the summary?

N/A – No meaningful error detected

5 – Error present, but no effect on overall understanding or usefulness

4 – Error could cause very minor confusion or misunderstanding

3 – Error could cause moderate confusion or loss of important nuance

2 – Error could cause major misunderstanding of key information

1 – Error changes the information so significantly that it could be harmful or highly misleading

## APPENDIX E Technology Acceptance Model - Perceived Ease of Use (TAM-PEU) Questionnaire

With 1 being strongly disagree to 7 being strongly agree, please indicate how much you agree or disagree with each statement about your experience using PCP-Bot.

- Learning to interact with PCP-Bot would be easy for me
- I would find it easy to get PCP-Bot to do what I want it to do
- My interaction with PCP-Bot would be clear and understandable
- I would find PCP-Bot to be flexible to interact with
- It would be essay for me to become skillful at using PCP-Bot
- I would find PCP-Bot easy to use

## APPENDIX F Adapted Human-Computer Trust Scale (HCTS) Questionnaire

Risk Perception

**RP1:** I believe that there could be negative consequences from using PCP-Bot

**RP2:** I feel I must be cautious when using PCP-Bot

**RP3:** It is risky to interact with PCP-Bot

**RP4:** I feel unsafe when interacting with PCP-Bot

**RP5:** I feel vulnerable when I interact with PCP-Bot

Competency

**COM1:** I believe PCP-Bot is competent and effective in collecting my medical and social history information

**COM2:** I believe PCP-Bot has the functionalities. I would expect from a medical history-taking chatbot

**COM3:** I believe PCP-Bot performs its role well in gathering my information for a physician’s review

Reciprocity

**REC1:** When I share information with PCP-Bot, I expect the follow-up to be relevant to what I said

**REC2:** When I provide information to PCP-Bot, I believe that I will get an appropriate follow-up

Benevolence

**BEN1:** I believe PCP-Bot acts in my best interest

**BEN2:** I believe PCP-Bot would do its best to help me during this medical history-taking process

**BEN3:** I believe PCP-Bot is designed to understand my needs and preferences during this conversation

General Trust

**GT1:** When I use PCP-Bot, I feel I can depend on it completely to collect my information accurately

**GT2:** I can always rely on PCP-Bot to follow the intended conversation flow

**GT3:** I can trust PCP-Bot to process and handle my responses appropriately for the physician’s use

## APPENDIX G NASA-TLX Questionnaire

With -10 being very low to +10 being very high, please indicate how much you agree or disagree with each statement about your experience using PCP-Bot.

- How mentally demanding was the conversation?
- How physically demanding was the conversation?
- How hurried or rushed was the pace of the conversation?
- How successful were you in accomplishing what you were asked to do?
- How hard did you have to work to accomplish your level of performance?
- How insecure, discouraged, irritated, stressed, and annoyed were you?

## Notes

### Competing Interest Statement

The authors have declared no competing interest.

### Funding Statement

National Library of Medicine and the NIH Office of the Director
project number 5DP1LM014558-03 (Former Number: 1DP1OD035237-01)

### Author Declarations

The synthetic patient cases used in this study were generated using an LLM (GPT-4.o). Txt files of those cases are located here: https://github.com/patrchen/PCP-Bot-Translator-Study-Cases This study was reviewed and deemed exempt by the Institutional Review Board of the University of Pennsylvania Human Research Protections Program (HRPP).

### Summary of Updates

Section 5 of Methods added to reflect funding declaration Table 4 of Results updated and revised

## References

1. Smeulers, M., Dikmans, M. & van Vugt, M. Well-prepared outpatient visits satisfy patient and physican. BMJ Open Qual. 8, e000496 (2019).

2. Tierney, A. A. et al. Ambient Artificial Intelligence Scribes to Alleviate the Burden of Clinical Documentation. NEJM Catal. 5, CAT.23.0404 (2024).

3. Lee, C., Britto, S. & Diwan, K. Evaluating the Impact of Artificial Intelligence (AI) on Clinical Documentation Efficiency and Accuracy Across Clinical Settings: A Scoping Review. Cureus 16, e73994.

4. Wong, M. et al. Addressing language barriers in U.S. healthcare: The role of CLAS standards, telehealth, and policy in supporting limited english proficiency populations. Health Sci. Rev. 17, 100249 (2025).

5. Wong, M. et al. Addressing language barriers in U.S. healthcare: The role of CLAS standards, telehealth, and policy in supporting limited english proficiency populations. Health Sci. Rev. 17, 100249 (2025).

6. Lindholm, M., Hargraves, J. L., Ferguson, W. J. & Reed, G. Professional language interpretation and inpatient length of stay and readmission rates. J. Gen. Intern. Med. 27, 1294–1299 (2012).

7. Masland, M. C., Lou, C. & Snowden, L. Use of Communication Technologies to Cost-Effectively Increase the Availability of Interpretation Services in Healthcare Settings. Telemed. J. E Health 16, 739–745 (2010).

8. Shool, S. et al. A systematic review of large language model (LLM) evaluations in clinical medicine. BMC Med. Inform. Decis. Mak. 25, 117 (2025).

9. Chow, J. C. L. & Li, K. Large Language Models in Medical Chatbots: Opportunities, Challenges, and the Need to Address AI Risks. Information 16, 549 (2025).

10. Sun, Z., Yang, J., Zhang, N., Wu, H. & Li, C. GPT-4o is more like a real person: potentials in surgical oncology. Int. J. Surg. Lond. Engl. 111, 1654–1655 (2024).

11. Menezes, M. C. S. et al. The potential of Generative Pre-trained Transformer 4 (GPT-4) to analyse medical notes in three different languages: a retrospective model-evaluation study. Lancet Digit. Health 7, e35–e43 (2025).

12. Yang, Y. et al. Beyond Multiple-Choice Accuracy: Real-World Challenges of Implementing Large Language Models in Healthcare. Annu. Rev. Biomed. Data Sci. 8, 305–316 (2025).

13. Ananda Rao, A., Chen, P.-L., Pugh, S. & Johnson, K. Developing an LLM-Based Conversational Agent for Primary-Care Pre-Visit Planning (PCP-Bot). (2025). doi:10.13140/RG.2.2.20413.17125.

14. Raymond, R. The 4 Most Common Languages Spoken Around the World | Community Ed at PCC. https://www.pcc.edu/community/2019/06/19/the-4-most-common-languages-spoken-around-the-world/.

15. Marian, V., Blumenfeld, H. K. & Kaushanskaya, M. The Language Experience and Proficiency Questionnaire (LEAP-Q): Assessing Language Profiles in Bilinguals and Multilinguals. J. Speech Lang. Hear. Res. 50, 940–967 (2007).

16. Kaushanskaya, M., Blumenfeld, H. K. & Marian, V. The Language Experience and Proficiency Questionnaire (LEAP-Q): Ten years later. Biling. Lang. Cogn. 23, 945–950 (2020).

17. Borsci, S., Schmettow, M., Malizia, A., Chamberlain, A. & van der Velde, F. A confirmatory factorial analysis of the Chatbot Usability Scale: a multilanguage validation. Pers. Ubiquitous Comput. 27, 317–330 (2023).

18. Borsci, S. & Schmettow, M. Re-examining the chatBot Usability Scale (BUS-11) to assess user experience with customer relationship management chatbots. Pers. Ubiquitous Comput. 28, 1033–1044 (2024).

19. Khanna, R. R. et al. Performance of an online translation tool when applied to patient educational material. J. Hosp. Med. 6, 519–525 (2011).

20. Technology Acceptance Model (TAM). Technology Acceptance Lab https://acceptancelab.com/technology-acceptance-model-tam.

21. Lee, A. T., Ramasamy, R. K. & Subbarao, A. Understanding Psychosocial Barriers to Healthcare Technology Adoption: A Review of TAM Technology Acceptance Model and Unified Theory of Acceptance and Use of Technology and UTAUT Frameworks. Healthcare 13, 250 (2025).

22. Beltrão, G., Sousa, S. & Lamas, D. Assessing the Measurement Invariance of the Human–Computer Trust Scale. Electronics 14, 1806 (2025).

23. Sousa, S. & Kalju, T. Modeling Trust in COVID-19 Contact-Tracing Apps Using the Human-Computer Trust Scale: Online Survey Study. JMIR Hum. Factors 9, e33951 (2022).

24. Hart, S. G. & Staveland, L. E. Development of NASA-TLX (Task Load Index): Results of Empirical and Theoretical Research. in Advances in Psychology (eds Hancock, P. A. & Meshkati, N.) vol. 52 139–183 (North-Holland, 1988).

25. Martínez, G. et al. Spanish is not just one: A dataset of Spanish dialect recognition for LLMs. Data Brief 63, 112088 (2025).

26. Mayor-Rocher, M. et al. It’s the same but not the same: Do LLMs distinguish Spanish varieties? Preprint at 10.48550/arXiv.2504.20049 (2025).

27. Bifulco, L. et al. A qualitative assessment of factors contributing to Spanish-speaking federally qualified health center patients’ chronic pain experiences. PLOS ONE 18, e0285157 (2023).

28. Cultures and Organizations: Software of the Mind, Third Edition.

29. Dang, Q. & Li, G. Unveiling trust in AI: the interplay of antecedents, consequences, and cultural dynamics. AI Soc. 41, 669–692 (2026).

